# Urinary viral shedding of COVID-19 and its clinical associations: A Systematic Review and Meta-analysis of Observational Studies

**DOI:** 10.1101/2020.05.15.20094920

**Authors:** Amir H Kashi, Jean de la Rosette, Erfan Amini, Hamidreza Abdi, Morteza Fallah-karkan, Maryam Vaezjalali

**Affiliations:** Urology and Nephrology Research Center (UNRC), Shahid Labbafinejad Hospital, Shahid Beheshti University of Medical Sciences, Tehran, Iran.; Department of Urology, Faculty of Medicine, Istanbul Medipol University, Istanbul, Turkey; Uro-Oncology Research Center, Tehran University of Medical Sciences, Tehran, Iran; Division of Urology, Department of Surgery, University of Ottawa, Ottawa, Canada; Department of Urology, Shohada Tajrish Hospital, Shahid Beheshti University of Medical Sciences (SBMU), Tehran, Iran.; Department of Microbiology, School of Medicine, Shahid Beheshti University of Medical Sciences, Tehran, Iran.

**Author notes:** Address for correspondence:*Maryam Vaezjalali, Department of Microbiology, School of Medicine, Shahid Beheshti University of Medical Sciences, Tehran, Iran.

**Keywords:** COVID-19, SARS-Cov-2, urine, review, meta-analysis, infection transmission

## Abstract

**Objectives:** To review the current literature on the presence of COVID-19 virus in the urine of infected patients and to explore the clinical features that can predict the presence of COVID-19 in urine.

**Materials and Methods:** A systematic review of published literature between 30^th^ December 2019 and 21^st^ June 2020 was conducted on Pubmed, Google Scholar, Ovid, Scopus, and ISI web of science. Studies investigating urinary viral shedding of COVID-19 in infected patients were included. Two reviewers selected relative studies and performed quality assessment of individual studies. Meta-analysis was performed on the pooled case reports and cohort with a sample size of ≥ 9.

**Results:** Thirty-nine studies were finally included in the systematic review; 12 case reports, 26 case series, and one cohort study. Urinary samples from 533 patients were investigated. Fourteen studies reported the presence of COVID-19 in the urinary samples from 24 patients. The crude overall rate of COVID-19 detection in urinary samples was 4.5%. Considering case series and cohorts with a sample size of ≥ 9, the estimated viral shedding frequency was 1.18 % (CI 95%: 0.14 – 2.87) in the meta-analysis. In adult patients, urinary shedding of COVID-19 was commonly detected in patients with moderate to severe disease (16 adult patients with moderate or severe disease versus two adult patients with mild disease). In children, urinary viral shedding of COVID-19 was reported in 4 children who all suffered from mild disease. Urinary viral shedding of COVID-19 was detected from day 1 to day 52 after disease onset. The pathogenicity of virus isolated from urine has been demonstrated in cell culture media in one study while another study failed to reveal replication of isolated viral RNA in cell cultures. Urinary symptoms were not attributed to urinary viral shedding.

**Conclusions:** While COVID-19 is rarely detected in urine of infected individuals, infection transmission through urine still remains possible. In adult patients, infected urine is more likely in the presence of moderate or severe disease. Therefore, caution should be exerted when dealing with COVID-19 infected patients during medical interventions like endoscopy and urethral catheterization.

## INTRODUCTION

Novel coronavirus disease (COVID-19), first reported from Wuhan, is a new disease caused by Severe Acute Respiratory Syndrome- Coronavirus-2 (SARS-CoV-2), manifesting mainly as an acute respiratory illness; however, the involvement of multiple organs including kidney and liver has been reported(1). The pathophysiological mechanisms for SARS-CoV-2 infection and organ invasion are still under investigation which leads to difficulties in understanding the routes of transmission, clinical diagnosis and treatment(2).

Genomic sequence analysis indicated that SARS-CoV-2 has almost 80% genomic similarity to coronavirus causing severe acute respiratory syndrome (SARS) namely SARS-CoV (3). In previous reports of SARS and the Middle East Respiratory Syndrome Coronavirus (MERS-CoV) infections, acute kidney injury was observed in 5% to 15% cases and was associated with a high (60%–90%) mortality rate(4).

The angiotensin-converting enzyme 2 (ACE2), known to be a cell receptor for human SARS-CoV, is also reported to play the same role for cellular entry of SARS-CoV-2 (5). In addition to respiratory organs, upregulation of ACE2 expression was also identified in urogenital system including renal proximal convoluted tubes, bladder urothelial cells (6) and genital organs including testis(7, 8).

The widely accepted routes of human to human transmission for COVID-19 are through respiratory droplets and direct contact; however, viral shedding in the urine has been reported and infection transmission through infected urine remains a possibility. The idea of virus transmission thorough urine originated from the homogeneity of the viral SARS-CoV-2 genome with the SARS virus and previous reports on the presence of SARS virus in urine(9, 10).

Original protocols for sample collections from COVID-19 patients included urine sample collection which further supports the likelihood of urine transmission in theory despite the fact that the mechanism of viral shedding is unclear(11). Two possible mechanisms for SARS-CoV-2 shedding in urine have been suggested: sepsis and cytokine storm results in renal dysfunction and subsequent leakage of SARS-CoV-2 from the circulation into urine; the virus may directly invade the urinary system via binding to ACE2 receptors and shed into the urine (12).

Although the viral shedding into the urine is hypothetically possible, most studies indicated that virus is absent in the urine of infected patients (6, 13-15). Conversely contradicting results come from reports confirming viral shedding in urine (1, 16, 17). It is therefore important to clarify whether viral transmission is possible through urine when it comes to manipulating the urinary system during endoscopic procedures. In line with this, clinicians will be guided to choose appropriate Personal Protecting Equipment (PPE) in preparation of endoscopic procedures. Likewise medical care workers will be able to take appropriate measures when handling urine samples or related procedures. It is also important to explore the clinical correlates of patients in whom viral shedding is observed to be able to better stratify patients into high and low risk of urinary viral shedding.

Considering the vast difference between reported viral shedding of COVID-19 in urine of infected patients in different series and lack of clinical correlates of urinary viral shedding in most reports, we performed a systematic review on the published literature to provide a summary estimate of the risk of COVID-19 infection from urine and to explore the clinical correlates of COVID-19 urinary shedding.

## MATERIALS AND METHODS

### Search strategy and data sources

We conducted a comprehensive systematic literature review of online databases, including Web of Science, PubMed, Scopus, Ovid, and Google Scholar from 1^st^ December 2019 till 21^st^ June 2020. Google scholar engine was set to search for every type of document. The search was performed by two independent investigators. The search terms used were: “(covid-19 OR ncovid-19 OR sars-cov-2 OR covid OR ncovid) AND urine”. The PICO terms for this review are: (P)atients are individuals infected with COVID-19; (I)ntervention is measurement of urinary viral shedding of COVID-19; and the (O)utcome would be determination of the frequency of COVID-19 urinary shedding in infected individuals.

Database searching was started on March 29^th^ 2020 and was regularly updated during extraction and analysis of retrieved studies to find newly published articles. The latest electronic search on cited databases was performed on June 21^st^, 2020.

References of retrieved articles were manually searched to find eligible studies. The search and selection criteria were restricted to English language.

### Study selection

The title and abstract of retrieved studies were screened through two different researchers (AHK, EA) independently. After removing duplicates and irrelevant studies, the full text of articles was examined for presence of original data on the presence of COVID-19 in urine. Any disagreement was resolved by a third person (MV). Personal viewpoints, opinion articles, correspondence, and letters not presenting original data were excluded as well as studies which did not report their result of urinary testing for COVID-19. Locations of studies was noted to identify duplicate case reports/series from the same area. For reports from the same area or possible reports from the same population of patients, the authors were contacted to provide clarification. The study protocol and reporting were evaluated in accordance with the Meta-Analysis of Observational Studies in Epidemiology (MOOSE) guidelines statement(18).

### Data Extraction

The main outcome in this study is the evaluation of viral shedding into the urine of patients infected with COVID-19 and clinical characteristics of patients in whom urinary viral shedding were reported. Data were extracted from the eligible manuscripts into pre-defined data-fields including study location, sample size, mean or median age, gender of patients, illness category, total number of patients and/or urine samples tested, urine assessment technique, total number of positive urine samples, and sampling time.

### Quality assessment of included studies

The included case series and cohort studies were evaluated in terms of quality according to the quality assessment tool for case series reported by the National Heart, Lung, and Blood Institute from the National Institutes of Health(19). This tool evaluates the quality based on a 9 item questionnaire. The questions focus on study population description, case definition, methods of including cases, comparability of included cases, description of interventions or assessments, follow-up and statistical methods used. Case reports were also evaluated using a similarly constructed checklist proposed by Murad et al. (20)

### Statistical methods

As case reports and case series with small sample size can suffer from selective reporting, they were excluded from the meta-analysis to provide an average estimate of urinary viral shedding for infected COVID-19 patients. Seventeen case series and one cohort study with a sample size of 9 cases or higher were included in the meta-analysis.

The effect size of individual studies was calculated by weighting each one of them by its inverse variance, and a confidence interval (CI) was thus obtained(21). Each study was weighted inversely proportional to its variance. To calculate the variance of each study, a binomial distribution was used. To investigate heterogeneity, the Q statistics and I2 index with *α* significance level of less than 10% were used. In this study, the random-effects model was considered, when there is heterogeneity among the studies (I2> 50%). The authors used the Egger’s test to check publication bias. Metaprop command in STATA used to stabilize the variances(22). STATA software (version 16) was used to analyze the data.

The relationship between disease severity in each study and frequency of viral shedding in the urine was investigated by weighting each study according to its sample size and performing spearman correlation.

## RESULTS

A total of 1238 articles were retrieved using the search strategy mainly through Google Scholar search engine. After studying the title and abstract of studies, and removing duplicate studies, the number was reduced to 179 studies. Full text of these 179 studies were studied and non-original studies such as communications without original data, personal reviews and letters were excluded resulting in 39 articles (Figure 1).

**Figure 1.**
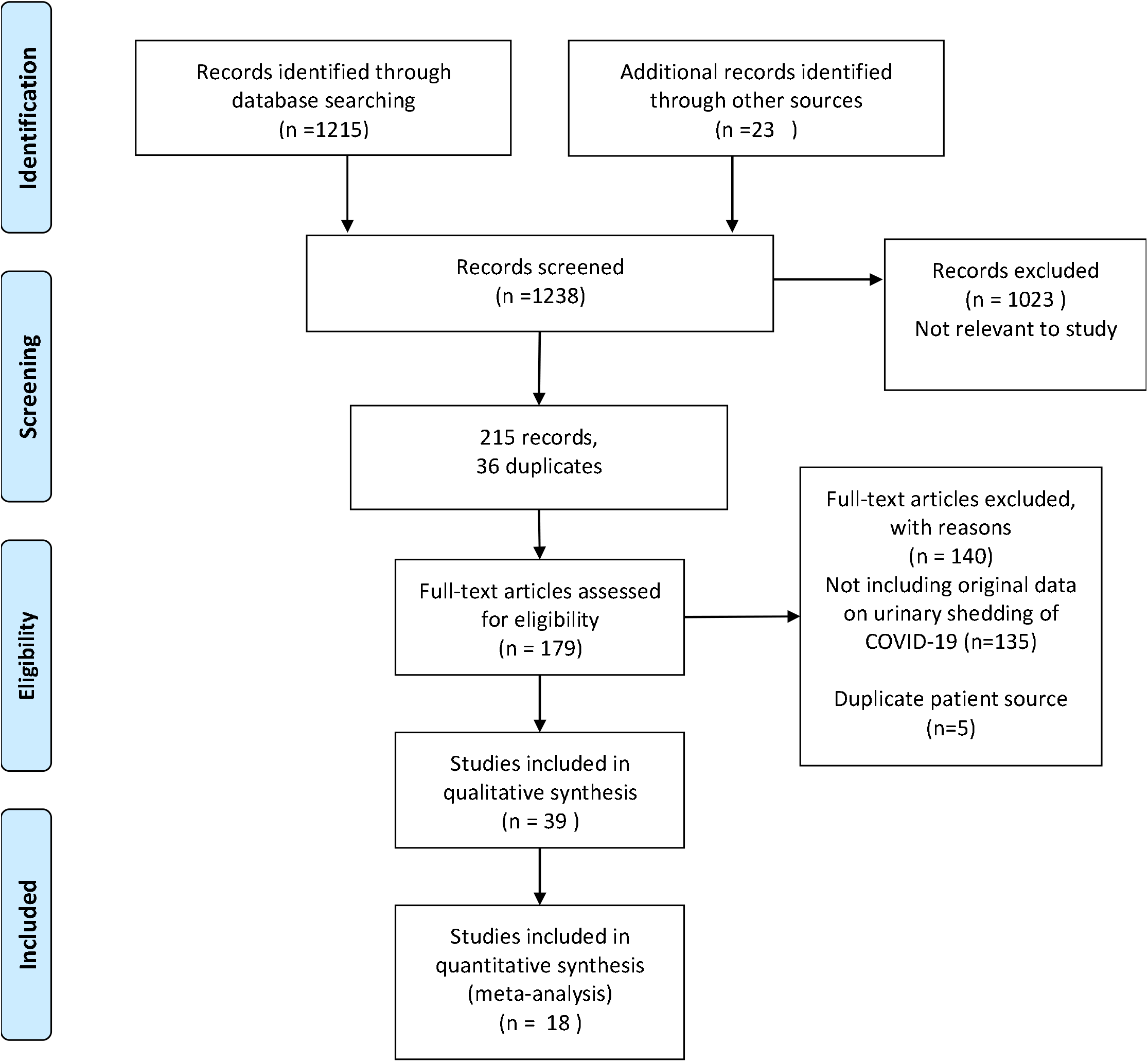
Flow diagram of included studies.

There were 12 case reports, 26 case series, and one cohort study from 12 different countries. The characteristics of the included studies are shown in Table 1. Out of the total reported population of 1995 patients in these 39 studies, urinary testing had been performed on 533 patients during admission and up to day 52 after illness onset. Positivity of urinary specimen was reported in 14 studies ranging from 1 to 4 positive urinary samples in each study summing to a total of 24 patients (4.5% frequency of viral shedding in patients’ urine). Positive urinary samples were reported from China (9 studies, 15 patients), Korea (4 studies, 7 patients), and Japan (1 study, 2 patients). The time of urine sampling in positive patients were reported as on admission day, day 1, day 3, day 5, day 7, day 9, day 10,, days 7-11, days 6 through 17, days 9 through 12, day 30, and day 52 after illness onset. In patient with positive urine sample on day 52, only urine sediment was positive and routine urine sample was negative in quantitative PCR for COVID-19.

**Table 1.**
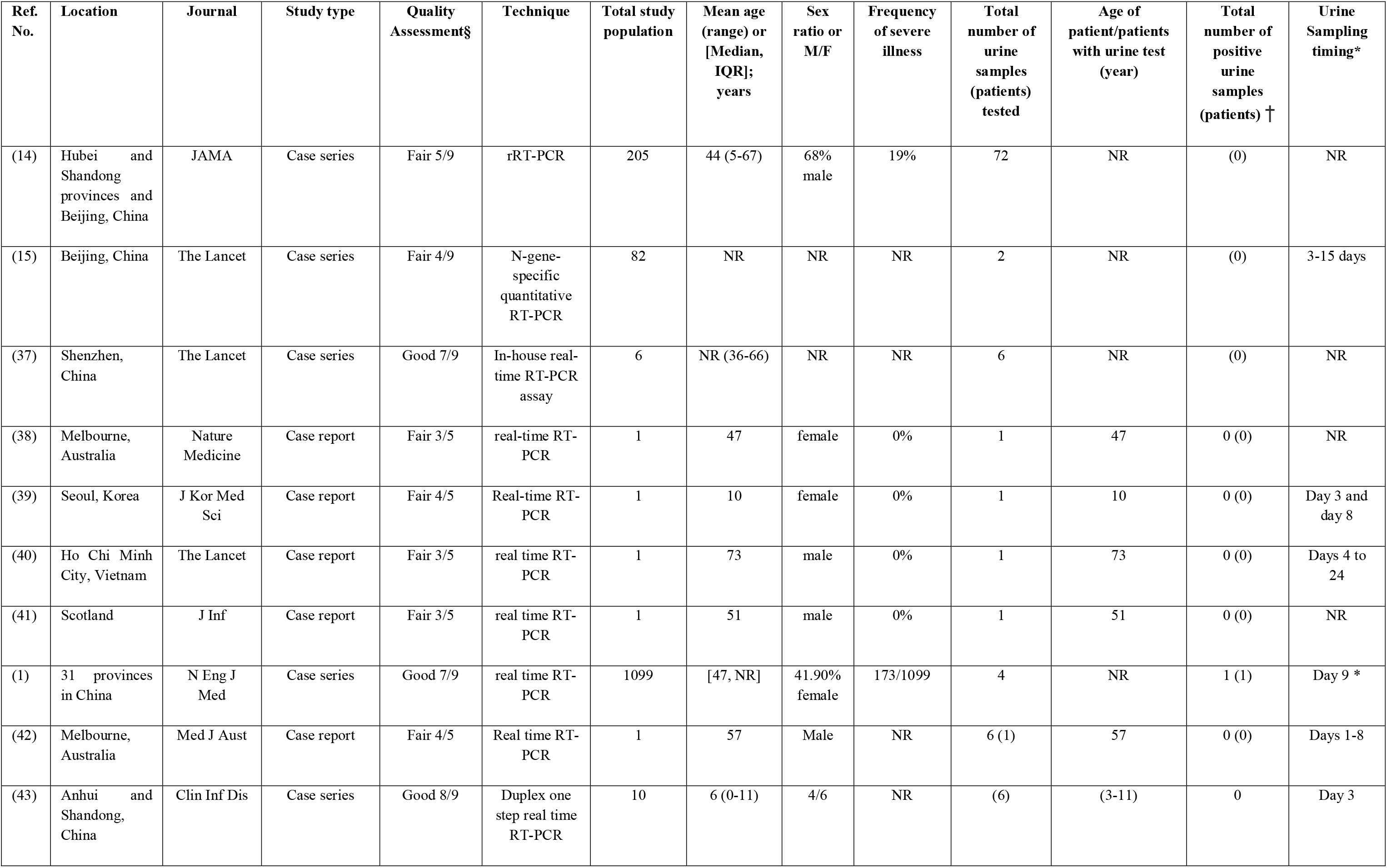
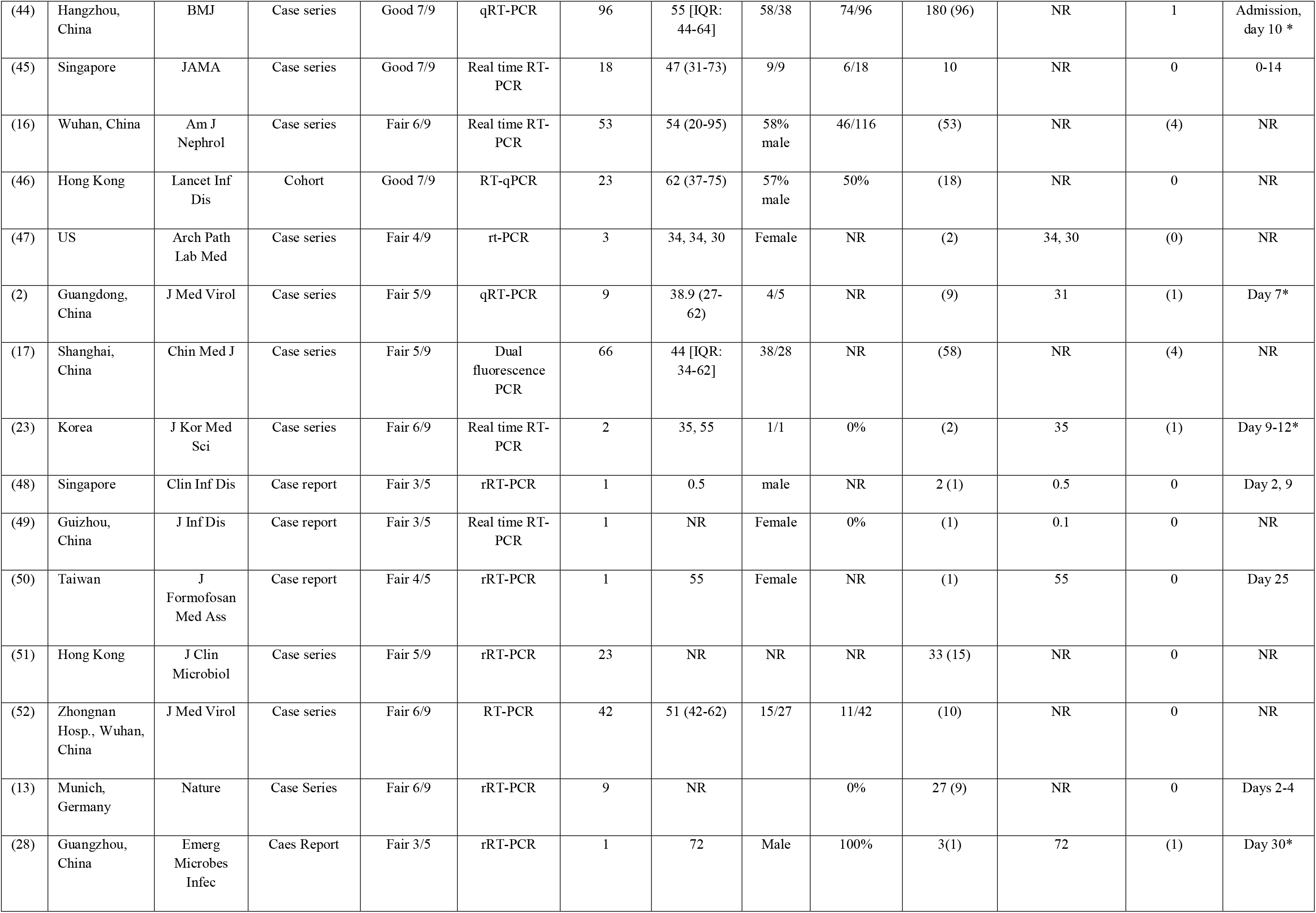
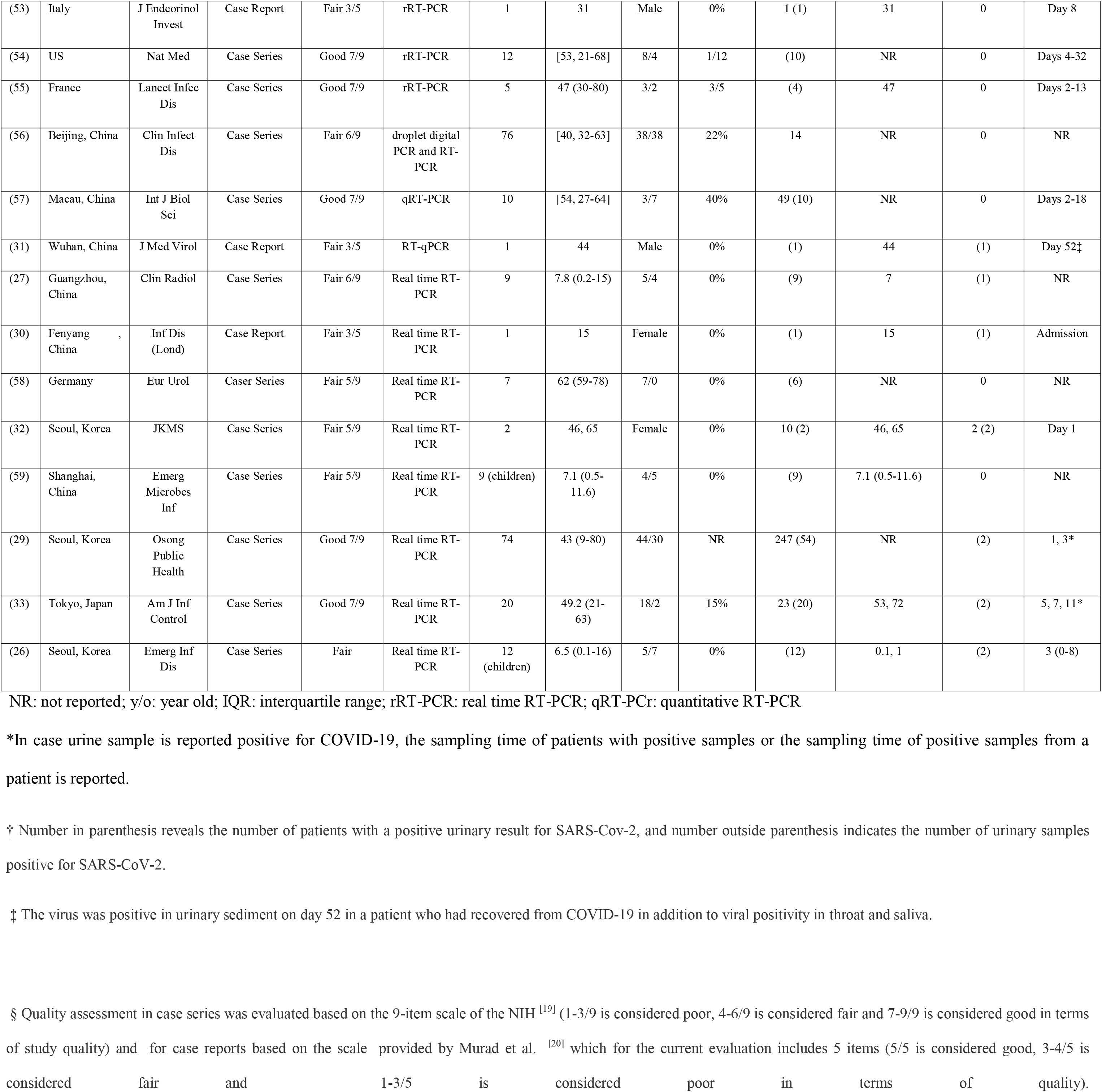
Characteristics of included studies.

| Ref. No. | Location | Journal | Study type | Quality Assessment§ | Technique | Total study population | Mean age (range) or [Median, IQR]; years | Sex ratio or M/F | Frequency of severe illness | Total number of urine samples (patients) tested | Age of patient/patients with urine test (year) | Total number of positive urine samples (patients) † | Urine Sampling timing* |
| --- | --- | --- | --- | --- | --- | --- | --- | --- | --- | --- | --- | --- | --- |
| (14) | Hubei and Shandong provinces and Beijing, China | JAMA | Case series | Fair 5/9 | rRT-PCR | 205 | 44 (5-67) | 68% male | 19% | 72 | NR | (0) | NR |
| (15) | Beijing, China | The Lancet | Case series | Fair 4/9 | N-gene-specific quantitative RT-PCR | 82 | NR | NR | NR | 2 | NR | (0) | 3-15 days |
| (37) | Shenzhen, China | The Lancet | Case series | Good 7/9 | In-house real-time RT-PCR assay | 6 | NR (36-66) | NR | NR | 6 | NR | (0) | NR |
| (38) | Melbourne, Australia | Nature Medicine | Case report | Fair 3/5 | real-time RT-PCR | 1 | 47 | female | 0% | 1 | 47 | 0 (0) | NR |
| (39) | Seoul, Korea | J Kor Med Sci | Case report | Fair 4/5 | Real-time RT-PCR | 1 | 10 | female | 0% | 1 | 10 | 0 (0) | Day 3 and day 8 |
| (40) | Ho Chi Minh City, Vietnam | The Lancet | Case report | Fair 3/5 | real time RT-PCR | 1 | 73 | male | 0% | 1 | 73 | 0 (0) | Days 4 to 24 |
| (41) | Scotland | J Inf | Case report | Fair 3/5 | real time RT-PCR | 1 | 51 | male | 0% | 1 | 51 | 0 (0) | NR |
| (1) | 31 provinces in China | N Eng J Med | Case series | Good 7/9 | real time RT-PCR | 1099 | [47, NR] | 41.90% female | 173/1099 | 4 | NR | 1 (1) | Day 9 * |
| (42) | Melbourne, Australia | Med J Aust | Case report | Fair 4/5 | Real time RT-PCR | 1 | 57 | Male | NR | 6 (1) | 57 | 0 (0) | Days 1-8 |
| (43) | Anhui and Shandong, China | Clin Inf Dis | Case series | Good 8/9 | Duplex one step real time RT-PCR | 10 | 6 (0-11) | 4/6 | NR | (6) | (3-11) | 0 | Day 3 |
| (44) | Hangzhou, China | BMJ | Case series | Good 7/9 | qRT-PCR | 96 | 55 [IQR: 44-64] | 58/38 | 74/96 | 180 (96) | NR | 1 | Admission, day 10 * |
| (45) | Singapore | JAMA | Case series | Good 7/9 | Real time RT-PCR | 18 | 47 (31-73) | 9/9 | 6/18 | 10 | NR | 0 | 0-14 |
| (16) | Wuhan, China | Am J Nephrol | Case series | Fair 6/9 | Real time RT-PCR | 53 | 54 (20-95) | 58% male | 46/116 | (53) | NR | (4) | NR |
| (46) | Hong Kong | Lancet Inf Dis | Cohort | Good 7/9 | RT-qPCR | 23 | 62 (37-75) | 57% male | 50% | (18) | NR | 0 | NR |
| (47) | US | Arch Path Lab Med | Case series | Fair 4/9 | rt-PCR | 3 | 34, 34, 30 | Female | NR | (2) | 34, 30 | (0) | NR |
| (2) | Guangdong, China | J Med Virol | Case series | Fair 5/9 | qRT-PCR | 9 | 38.9 (27-62) | 4/5 | NR | (9) | 31 | (1) | Day 7* |
| (17) | Shanghai, China | Chin Med J | Case series | Fair 5/9 | Dual fluorescence PCR | 66 | 44 [IQR: 34-62] | 38/28 | NR | (58) | NR | (4) | NR |
| (23) | Korea | J Kor Med Sci | Case series | Fair 6/9 | Real time RT-PCR | 2 | 35, 55 | 1/1 | 0% | (2) | 35 | (1) | Day 9-12* |
| (48) | Singapore | Clin Inf Dis | Case report | Fair 3/5 | rRT-PCR | 1 | 0.5 | male | NR | 2 (1) | 0.5 | 0 | Day 2, 9 |
| (49) | Guizhou, China | J Inf Dis | Case report | Fair 3/5 | Real time RT-PCR | 1 | NR | Female | 0% | (1) | 0.1 | 0 | NR |
| (50) | Taiwan | J Formosan Med Ass | Case report | Fair 4/5 | rRT-PCR | 1 | 55 | Female | NR | (1) | 55 | 0 | Day 25 |
| (51) | Hong Kong | J Clin Microbiol | Case series | Fair 5/9 | rRT-PCR | 23 | NR | NR | NR | 33 (15) | NR | 0 | NR |
| (52) | Zhongnan Hosp., Wuhan, China | J Med Virol | Case series | Fair 6/9 | RT-PCR | 42 | 51 (42-62) | 15/27 | 11/42 | (10) | NR | 0 | NR |
| (13) | Munich, Germany | Nature | Case Series | Fair 6/9 | rRT-PCR | 9 | NR |  | 0% | 27 (9) | NR | 0 | Days 2-4 |
| (28) | Guangzhou, China | Emerg Microbes Infec | Caes Report | Fair 3/5 | rRT-PCR | 1 | 72 | Male | 100% | 3(1) | 72 | (1) | Day 30* |
| (53) | Italy | J Endocrinol Invest | Case Report | Fair 3/5 | rRT-PCR | 1 | 31 | Male | 0% | 1 (1) | 31 | 0 | Day 8 |
| (54) | US | Nat Med | Case Series | Good 7/9 | rRT-PCR | 12 | [53, 21-68] | 8/4 | 1/12 | (10) | NR | 0 | Days 4-32 |
| (55) | France | Lancet Infect Dis | Case Series | Good 7/9 | rRT-PCR | 5 | 47 (30-80) | 3/2 | 3/5 | (4) | 47 | 0 | Days 2-13 |
| (56) | Beijing, China | Clin Infect Dis | Case Series | Fair 6/9 | droplet digital PCR and RT-PCR | 76 | [40, 32-63] | 38/38 | 22% | 14 | NR | 0 | NR |
| (57) | Macau, China | Int J Biol Sci | Case Series | Good 7/9 | qRT-PCR | 10 | [54, 27-64] | 3/7 | 40% | 49 (10) | NR | 0 | Days 2-18 |
| (31) | Wuhan, China | J Med Virol | Case Report | Fair 3/5 | RT-qPCR | 1 | 44 | Male | 0% | (1) | 44 | (1) | Day 52‡ |
| (27) | Guangzhou, China | Clin Radiol | Case Series | Fair 6/9 | Real time RT-PCR | 9 | 7.8 (0.2-15) | 5/4 | 0% | (9) | 7 | (1) | NR |
| (30) | Fenyang, China | Inf Dis (Lond) | Case Report | Fair 3/5 | Real time RT-PCR | 1 | 15 | Female | 0% | (1) | 15 | (1) | Admission |
| (58) | Germany | Eur Urol | Caser Series | Fair 5/9 | Real time RT-PCR | 7 | 62 (59-78) | 7/0 | 0% | (6) | NR | 0 | NR |
| (32) | Seoul, Korea | JKMS | Case Series | Fair 5/9 | Real time RT-PCR | 2 | 46, 65 | Female | 0% | 10 (2) | 46, 65 | 2 (2) | Day 1 |
| (59) | Shanghai, China | Emerg Microbes Inf | Case Series | Fair 5/9 | Real time RT-PCR | 9 (children) | 7.1 (0.5-11.6) | 4/5 | 0% | (9) | 7.1 (0.5-11.6) | 0 | NR |
| (29) | Seoul, Korea | Osong Public Health | Case Series | Good 7/9 | Real time RT-PCR | 74 | 43 (9-80) | 44/30 | NR | 247 (54) | NR | (2) | 1, 3* |
| (33) | Tokyo, Japan | Am J Inf Control | Case Series | Good 7/9 | Real time RT-PCR | 20 | 49.2 (21-63) | 18/2 | 15% | 23 (20) | 53, 72 | (2) | 5, 7, 11* |
| (26) | Seoul, Korea | Emerg Inf Dis | Case Series | Fair | Real time RT-PCR | 12 (children) | 6.5 (0.1-16) | 5/7 | 0% | (12) | 0.1, 1 | (2) | 3 (0-8) |

NR: not reported; y/o: year old; IQR: interquartile range; rRT-PCR: real time RT-PCR; qRT-PCR: quantitative RT-PCR
\*In case urine sample is reported positive for COVID-19, the sampling time of patients with positive samples or the sampling time of positive samples from a patient is reported.
† Number in parenthesis reveals the number of patients with a positive urinary result for SARS-Cov-2, and number outside parenthesis indicates the number of urinary samples positive for SARS-CoV-2.
‡ The virus was positive in urinary sediment on day 52 in a patient who had recovered from COVID-19 in addition to viral positivity in throat and saliva.
§ Quality assessment in case series was evaluated based on the 9-item scale of the NIH <sup>[19]</sup> (1-3/9 is considered poor, 4-6/9 is considered fair and 7-9/9 is considered good in terms of study quality) and for case reports based on the scale provided by Murad et al. <sup>[20]</sup> which for the current evaluation includes 5 items (5/5 is considered good, 3-4/5 is considered fair and 1-3/5 is considered poor in terms of quality).

As indicated above, a meta-analysis was performed on case series with urinary investigations in 9 or more patients. Fixed effect model was used as heterogeneity among studies was low (I^2^=18 %). The meta-analysis forest plot which takes into account the weight of each study according to the inverse of its variance revealed a pooled estimate of 1.18 % (CI 95%: 0.14 – 2.87%) for viral shedding in urine of patients (Figure 2). The Begg’s funnel plot revealed no publication bias (Figure 3).

**Figure 2.**
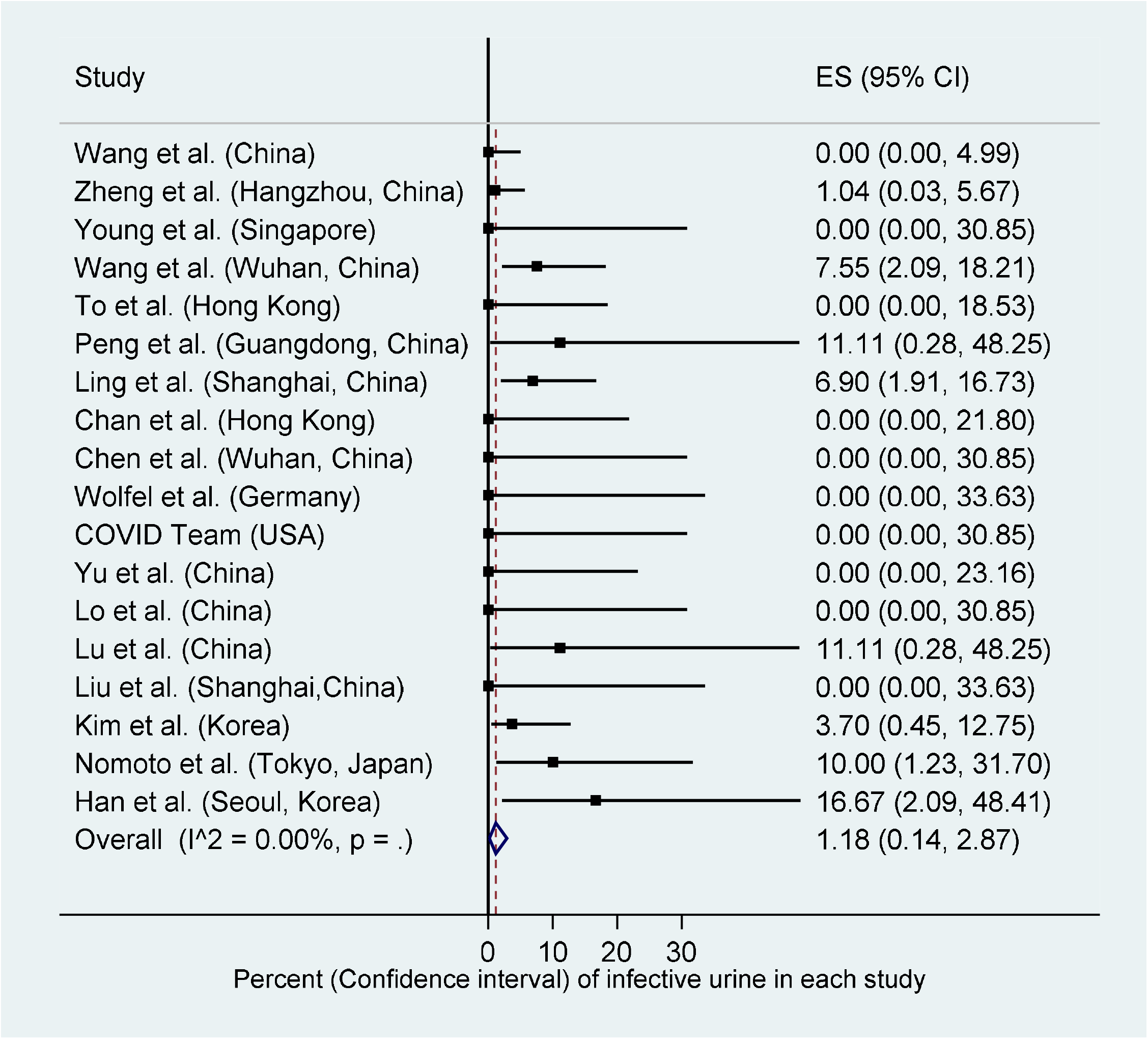
Forest plot of the frequency of urinary viral shedding in each study and the pooled estimate.

**Figure 3.**
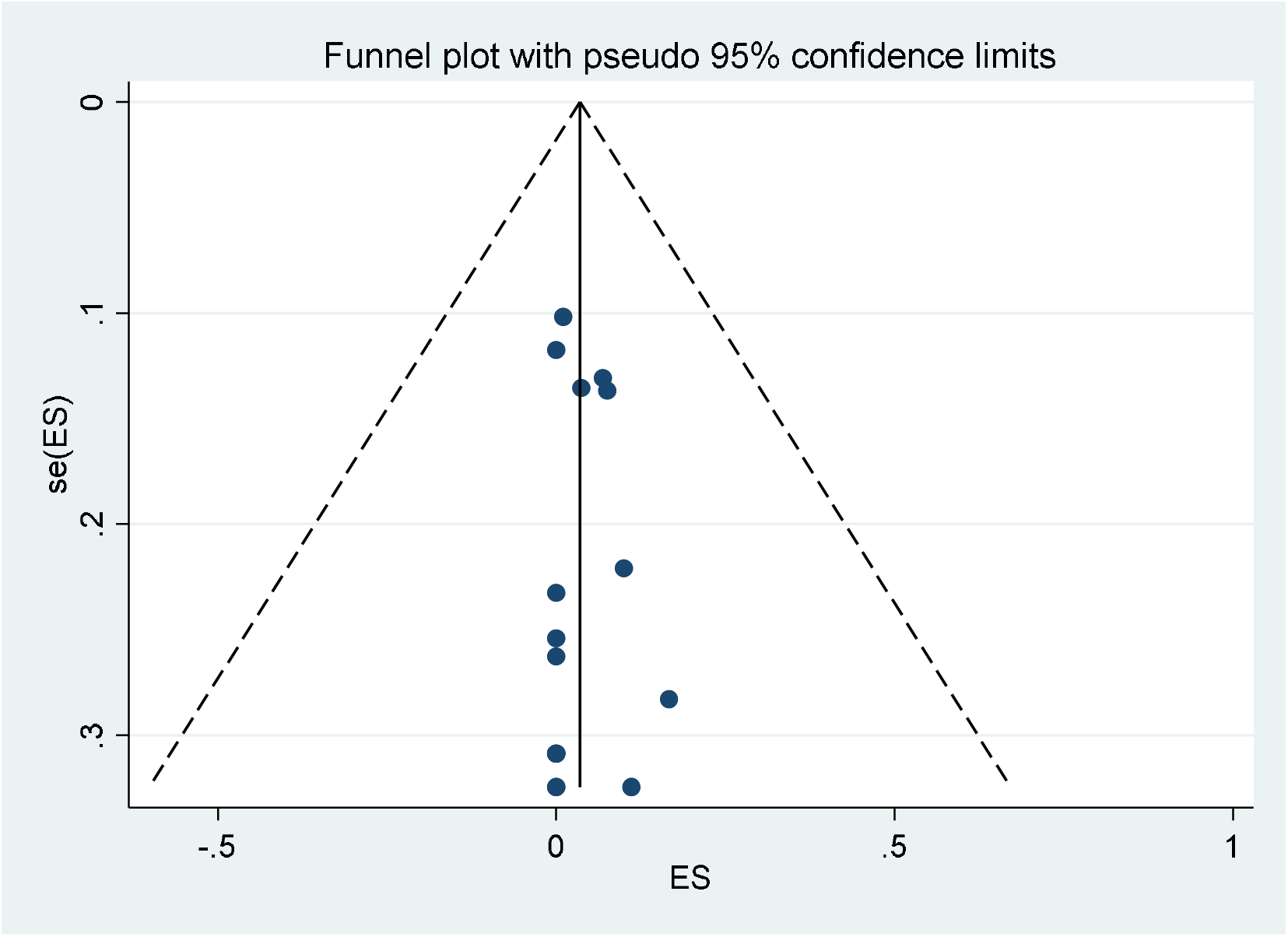
Funnel plot for evaluation of publication bias.

One of the studies which confirmed the presence of COVID-19 RNA in the urine stated that the degree of positivity of the urine did not meet the reference for positivity in rRT-PCR (real time reverse transcriptase PCR); however, the level of detected E and RdRp genes on days 9 and 12 post diagnosis were higher than cut off values and it remained positive until recovery. This patient’s urine sample was considered positive in the current review(23).

Peng et al. reported the presence of COVID-19 in urine of one of the 9 studied patients on day 7 after the symptom onset. The patient’s urine sample turned negative on day 10. In this study, the viral load in urine sample was lower than rectal and oropharyngeal samples. It is interesting to note that the patient with positive urinary PCR for COVID-19 did not complain of any urinary symptoms(24).

Wang et al. investigated urinary samples of patients with chronic kidney disease (CKD) versus patients with normal renal function. Urinary PCR was positive in one out of 5 patients with CKD versus 3 out of 48 patients with normal renal function. In this study, the clinical course and characteristics of patients with detected COVID-19 in the urine were not different from patients without it(16).

Ling and colleagues reported 66 patients with COVID-19 from Shanghai, China. Urine samples of 4 patients (6.9%) were positive for COVID-19. Interestingly, in 3 patients, urinary samples remained positive even after clearance of virus in oropharyngeal samples(17).

Han et al. reported the presence of COVID-19 virus in the urine of a newborn from an infected mother. The virus was discovered in samples from the oropharynx, saliva, urine and faeces. Despite the fact that the detected urinary viral load was relatively low, it was above the diagnostic cut off on days 6 through 17 after the onset of illness (11 days). Once again, the urinary viral load was still positive after clearance of virus from nasopharyngeal and plasma samples(25). In another publication, Han and colleagues investigated urinary viral shedding of children in Seoul, Korea. This report includes the data of the neonate indicated above with another one-year-old male infant. Urinary viral loads were 3.82, 7.55 log_10_ copies/mL. Urinary samplings were performed in median (range) days of 3 (0-8) after illness. Nine children were asymptomatic and 3 children suffered from mild symptoms (26).

In another study in children, the urine sample of 1 out of nine infected children was positive for COVID-19 by real time RT-PCR detection method. All children in this study were either asymptomatic or had mild disease. The urine was positive in a 7 year-old girl who presented only with fever (38.7 ° C) without any cough or respiratory symptoms (27).

In the communication of Sun and colleagues, the urine samples of a 72 year-old male with severe COVID-19 infection was investigated. Urinary sampling was performed on days 12, 30 and 42 after symptom onset. The viral load was above the diagnostic threshold only on day 30. The authors inoculated Vero E6 cells with urine of patient on day 12 (with viral load below the diagnostic threshold). Interestingly, cytopathic effects were observed after 3 days of inoculation. Electron microscopy revealed the presence of the virus in inoculated cells by demonstration of spherical-shaped particles with distinct surface projections, resembling spikes. The authors furthermore compared serum sample from this patient, who had high IgM and IgG against SARS-CoV-2, and a healthy control individual and demonstrated staining of inoculated cells in immunofluorescent assay only in the patient serum (28).

On the contrary, Kim et al. reported 2 infected urine samples from two patients out of 54 patients who were investigated by urinary testing for COVID-19. The virus was isolated on day 1 and 3 after admission in these two patients. Urinary viral loads were 49, and 109 copies/μL.

Subsequent testing in days 2-11 in the first patient and day 6 in the second patient failed to reveal urinary shedding of COVID-19. Subsequently, CaCo-2 cells were inoculated with infected urine. Viral RNA could not be isolated form infected cell culture after 5 days. Therefore, the authors suggested a low possibility for transmission of COVID-19 through urine (29).

Ren and colleagues reported a 15-year-old female who was admitted with mild COVID-19. The initial diagnosis of COVID-19 was made in her urine sample while her throat sample was negative for COVID-19 at that time. The throat sample turned positive for COVID-19 two days later. (30)

Yang and colleagues reported *urine sediment* positivity in a 44 year-old man who had initially recovered with confirming negative throat swab test but then relapsed with positive throat and salivary rRT-PCR results. The urine was negative for COVID-19 in RT-PCR on day 52 after illness onset;however, urinary sediment was positive for COVID-19 in RT-PCR on the same day (31).

Yoon and colleagues reported two positive urine samples from two infected female patients from Seoul, Korea. Urinary samples were both positive on the first day of admission and were negative when tested at days 3, 5, 7, and 9 after admission. The urinary viral loads on the admission day were 5.48 log _10_ and 5.79 log _10_ copies/mL (32).

Nomoto et al. reported urinary investigation of COVID-19 in 20 patients hospitalized in Tokyo, Japan. Two patients (one with moderate disease needing oxygen supplementation and another patient with severe disease needing ventilator support) were positive for urinary viral shedding of COVID-19. The urine of the patient with moderate disease severity, was positive only on day 5 after disease onset while the urine of patient with severe disease was positive on days 7 and 11 after disease onset. Urinary viral loads were 840, 800, and 254 copies/mL on days 5, 7, and 11 of illness as described above. The urinary viral shedding of COVID-19 was 0/9 in patients with mild disease, 1/8 in patients with moderate disease and 1/3 in patients with severe disease. The authors advised careful handling of urine in patients with moderate to severe disease (33).

The severity of COVID-19 was reported for 22 patients out of 24 patients with urinary viral shedding of COVID-19. In adult patients, 16 out of 18 patients with urinary vial shedding of COVID-19 suffered from moderate or severe disease. On the other hand, in children all 4 children with urinary viral shedding of COVID-19 suffered from mild illness (Table 1). The association of disease severity and frequency of urinary viral shedding in studies on adult patients with a sample size of 9 and over (9 studies) was statistically significant (spearman r=0.67, P < 0.001).

## DISCUSSION

This report provides a comprehensive overview of the available evidence from 30^th^ December 2019 to 21^st^ June 2020 on detection of SARS-CoV-2 in the urine samples. Thirty-nine studies from 12 countries were included with a total of 533 patients in whom results of urinary testing for COVID-19 were reported. Initially during the COVID-19 outbreak, evaluation and investigation of urinary samples were considered as part of routine sampling as stated by the World Health Organization interim guideline for laboratory testing for COVID-19(11). Later publications pointed to the rarity of viral presence in urine or totally rejected the presence of COVID-19 in urine (34). Recently, several publications reported the detection of COVID-19 in the urine which made us conduct this systematic review to find the likelihood of positive urine test in a COVID-19 patient.

The overall viral shedding of COVID-19 in urine of infected individuals was 4.5 % in 533 patients in whom urinary testing was done for COVID-19. Excluding case reports and case series with small sample size the estimated pooled frequency of urinary viral shedding was 01.18 % (CI 95%: 0.14 – 2.87) in the meta-analysis. Therefore one can conclude that the probability to detect the virus in urine is at least 1.18 %. In adult patients, urinary viral shedding was highly correlated with disease severity as only 11% (2/18) of adult patients with urinary viral shedding suffered from mild disease. In children this correlation was not observed as all children with urinary viral shedding of COVID-19 suffered from mild disease. As for urinary symptoms, no association has been reported between symptoms and presence of virus in urine (2). The presence of proteinuria and microscopic hematuria in severe disease(35) may be explained by cytokine storm ^(12)^ in a systemic disease rather than direct invasion of renal parenchyma and urinary tract and is most likely non-specific. These findings suggest that one cannot screen the patients either based on positive urinary symptoms which reiterates the need for consideration of precaution.

An important clinical question is the potential of viral RNA isolated form urine to infect other individuals. Real time RT-PCR for detection of COVID-19 measures the presence of viral genome particles in urine, however infectivity necessitates the presence of virion including envelope and capside. Two studies have tried to investigate the above concern (28, 29). Both studies aimed to investigate infectivity of urine by inoculating cell cultures by infected urine. Sun et al. confirmed cytopathic effects of urinary virus in Vero E6 cell culture by electron microscopy and immunofluroscent staining while Kim et al. failed to document replication of isolated virus in CaCo2 cell culture. Therefore, confirmation of urine infectivity needs further trials in future.

Another important clinical question is about the duration of necessary precautions in case we actually need it. There are reports indicating that the virus can be detected in the urine despite other negative test from different specimen (17, 25, 30). Therefore, a negative throat swab does not rule out the need for urinary screening in case urinary tract related procedures are planned. This can also be different in children with a reported longer viral shedding period in the urine (25) and will need future clarifications with better designed studies.

The highest frequency of infected urinary samples (in studies with nine or more patients) belongs to the reports of Han et al.(26), Peng et al.(24), and Lu et al.(27) from Seoul, Guangdong, and Guangzhou who reported 16.7% (2/12), 11.1% (1/9), and 11.1% (1/9) percent for infected urinary samples in their studies. But even this rate is greatly lower in comparison with urinary infection rates of SARS-CoV which was approximately 42%(36). One of the possible reasons for the low detection rate of COVID-19 can be the short duration of viral presence in the urine. Kim et al.(23) investigated urinary samples from two Korean patients from day 3 through day 14 after the onset of illness. The PCR for RdRp was marginally positive only on day 12 and for gene E, the PCR was again marginally positive only on day 9. Another cause can be low quantity of virus in urinary samples which makes its detection in real time PCR assays difficult. However, as indicated above Sun et al.(28) suggested the pathogenicity of low urinary viral load in cell cultures. This will raise another concern regarding the handling of the urinary specimen even with a negative urine test results and further need for using PPEs until better designed studies clarify the likelihood of disease transmission via urine.

Our study has some limitations. Given the nature of systematic reviews and meta-analysis our results rely on the original studies which are mostly observation studies. Despite the fact that this is a comprehensive review of current knowledge and sheds light on some unknown areas of uncertainty, we cannot definitely conclude if the virus transmits via urine and if so for how long the protection is needed. All we can say is that the virus is detectable with lower quantity in the urine of a smaller cohort of patients likely not proportionate to the clinical symptoms and the risk of transmission remains possible.

## CONCLUSIONS

The results of this review reveal an estimated positivity rate of 1.18% for COVID-19 in patients’ urine samples. Urinary viral load in most reports were lower than rectal or oropharyngeal samples. In adult patients, urinary viral shedding was more commonly observed in individuals with moderate or severe disease while in children viral shedding was also observed in patients with mild disease. Despite the fact that our findings reiterate the low detection rate of urinary COVID-19, based on the in vitro reports of potential transmission, we suggest regular caution in handling urinary samples in patients with COVID-19 infection and more importantly when performing procedural interventions such as urethral catheter insertion or endourologic interventions.

## Data Availability

All data of manuscript is available in case needed.

## DECLARATION OF INTERESTS

He authors report no conflict of interest.

## AUTHORS’ CONTRIBUTIONS

AHK concepted of the study, helped in data gathering, helped in data analysis, and drafted the article.

JR helped in data interpretation, drafting and revising the manuscript.

MF helped in data gathering and in revising the manuscript.

EA helped in data gathering and in manuscript revision.

HA contributed in data interpretation, manuscript draft and revision.

MV helped in data gathering, drafting and revising the manuscript.

All authors approved the final version of the manuscript.

## Notes

### Competing Interest Statement

The authors have declared no competing interest.

### Funding Statement

No funding.

### Author Declarations

This is a sys review and the ethics has been approved by the UNRC.

